# The association between self-reported vision and mental wellbeing: A secondary analysis of Health Survey for England data

**DOI:** 10.1101/2025.03.06.25323492

**Authors:** Michael D. Crossland, Tessa M. Dekker, Marc S. Tibber

## Abstract

**Objectives:** Eye disease and vision impairment are known to be associated with reduced mental wellbeing but less is known about the wellbeing of people with near-normal levels of vision. Here we examined the association between self-reported eyesight and mental wellbeing, controlling for eye disease, mental ill-health and demographic factors, for adults with a wide range of age and vision.

**Design:** Population-based cross-sectional study.

**Participants:** 7705 adults aged 16+ who participated in Health Survey for England 2013, self-reported their eyesight status and completed the Warwick Edinburgh Mental Wellbeing Scale.

**Primary outcome measure:** Mental wellbeing, controlling for the presence of self-reported mental ill health, self-reported eye disease, age, sex, socioeconomic group and ethnic origin.

**Results:** Poorer self-reported eyesight was strongly associated with lower mental wellbeing (*F* _(4, 7700)_ = 94.7, *p* < 0.0001). This association remained significant after controlling for self-reported mental ill health, self-reported eye disease, age, sex, socioeconomic group and ethnic origin.

**Conclusions:** Self-reported eyesight is strongly associated with mental wellbeing, irrespective of whether people have vision impairment or a diagnosed eye disease. This relationship exists in people with and without mental ill-health. Mental wellbeing should be considered in people with reduced eyesight, regardless of whether they have a diagnosed eye disease or mental ill-health. Interventions which improve vision may have a positive impact on mental wellbeing.

**Strengths and weaknesses:**

- This is the first study to investigate the relationship between eyesight and mental wellbeing in a large cohort of adults with a wide range of age and level of vision.
- We have corrected for multiple potentially confounding factors including mental ill-health, demographic and socioeconomic characteristics.
- We rely on self-report of eyesight, eye disease and mental ill-health.
- The cross-sectional nature of Health Survey for England means we are unable to determine causality, particularly whether reduced eyesight directly leads to reduced wellbeing, whether those with lower wellbeing underestimate their vision, or whether people with lower wellbeing experience poorer vision (perhaps due to differences in health behaviours)

**Funding statement:** This work was supported by the Macular Society (research grant 22-RG-1) and by the National Institute for Health and Care Research (NIHR) Biomedical Research Centre at Moorfields Eye Hospital NHS Foundation Trust and UCL Institute of Ophthalmology. The views expressed are those of the author(s) and not necessarily those of the NHS, the NIHR or the Department of Health and Social Care.

## Introduction

Almost two million people in the UK have vision impairment,[1] of whom about 134,000 are registered as sight impaired (formerly called ‘partially sighted’) and around the same number are registered *severely* sight impaired (previously ‘blind’) [2]. People with vision impairment experience poorer mental wellbeing [3], lower life satisfaction [4] and increased psychological distress [5] relative to those without. In a longitudinal study of older adults, Xiang and colleagues found that vision had a stronger association than dementia on wellbeing, but that depression had a even greater role on wellbeing.[3]

Many people have reduced eyesight yet do not meet the World Health Organisation’s definition of vision impairment (broadly speaking, those with visual acuity less than 6/12 in their better eye, or with significant visual field loss). This can be due to mild eye disease (such as early cataract), uncorrected refractive error, amblyopia (often caused by suboptimal eye care in early life) or by not having access to the optimal spectacle correction. In a large UK-based population study, only 77% of adults were found to have ‘normal’ vision in both eyes with their current spectacles or contact lenses (if worn).[6]

The impact of this subthreshold vision impairment on wellbeing is not well understood, despite the implications this might have for treatment and intervention. For example, if early vision impairment was associated with poorer wellbeing, this would encourage earlier treatment for diseases like cataract. One study has shown a relationship between poorer self-reported vision, tearfulness, lack of enjoyment, hopelessness, difficulty concentrating and other aspects of wellbeing.[7] However, this study only investigated older adults, did not use a standardised measure of mental wellbeing, and did not correct for the higher levels of depression also found in people with poorer levels of eyesight.

The interaction between self-reported vision and wellbeing is complicated by the well-established links between eye disease, depression [8] and anxiety.[9] For example, the odds of experiencing depression is approximately doubled in people with vision impairment[10]. However, this is potentially confounded by activity limitation (a common ‘symptom’ of depression), which can be incorrectly attributed to depression, yet actually caused by vision impairment: somebody may respond ‘yes’ to the screening question ‘have you dropped many of your activities and interests?’ not because they are depressed, but because they are no longer able to see well enough to continue their hobbies.[11] It is unclear whether the lower wellbeing experienced by people with vision impairment is simply caused by the increased level of depression and mental-ill health in this population, or whether vision impairment has an independent impact on wellbeing.

The interaction between vision impairment and wellbeing is further complicated by demographic, health and societal factors, many of which are associated with an increased likelihood of vision impairment and eye disease, as well as mental ill-health and poor wellbeing.[6,12–17] For example, experience of childhood poverty has been linked to increased rates of a range of mental health difficulties, including depression,[18,19] as well as physical health difficulties, including low vision.[14,20]

In this study, we performed a secondary analysis on a large dataset of adults with a wide age range, to examine the relationship between eyesight and mental wellbeing while controlling for mental ill-health, demographic and socioeconomic characteristics. Our primary hypothesis (H1) was that reduced vision would be associated with poorer wellbeing. Our secondary hypotheses (H2) were that this effect would survive after controlling for (i) mental ill-health, (ii) eye disease, and (iii) demographic and socioeconomic factors. Confirmation of these secondary hypotheses would have potential implications for the ophthalmological treatment of people with early eye disease, and on the provision of interventions to improve wellbeing in people reporting reduced vision.

## Method

A secondary analysis of data collected as part of the 2013 Health Survey for England (HSE) was undertaken. HSE has been administered annually since 1991 [21] and since 2010 has included a self-reported assessment of mental wellbeing. In 2013, HSE additionally included items on vision, including self-reported eyesight when wearing spectacles or contact lenses and the presence of diagnosed eye disease.

### Participant identification

For the original study, participants were identified using a random selection of 9,408 addresses in 588 postcode sectors across England. Every adult and up to two of the children in each selected household was eligible to participate. Data were collected in two stages. In the first, each participant completed an interview undertaken by a researcher and was given a self-completion booklet. The second involved a nurse visit, during which additional questions were asked and a blood sample was taken.

### Demographic details

Age at last birthday and ethnic group were self-reported. Household socio-economic group was coded by the original study team, by deriving the National Statistics Socio-economic Classification (NS-SEC)[22] from the occupation of a ‘reference person’ within the household. The reference person was defined as the person in whose name the property was owned or rented; if there was more than one, the person with the highest income was used. If there were two householders with equal income, then the oldest person was chosen as household reference person.[21] For our analyses, the NS-SEC3 three-category scale was used, where socio-economic status is classified as: (i) higher managerial/professional occupations, (ii) intermediate occupations, (iii) routine and manual occupations, with an option to report (iv) other.

### Level of vision and eye disease

In the first stage of data collection for the 2013 HSE, participants were asked ‘Using glasses or corrective lenses if you use them, is your eyesight excellent, very good, good, fair or poor?’.

Participants were also asked whether they had any ‘health conditions, illnesses or impairments, lasting or expected to last 12 months or more’ and were able to list up to six conditions. These conditions were coded by the interviewer. Participants were identified as having eye disease when the interviewer coded ‘cataract/poor eye sight/blindness’ or ‘other eye complaints’.

Additionally, participants were asked whether a doctor or optician had ever told them they had macular degeneration, cataract (and, if so, whether they had had surgery), diabetic eye disease or diabetic retinopathy, glaucoma or suspected glaucoma, injury or trauma resulting in loss of vision, or another serious eye condition; and if they were registered as ‘blind or partially sighted’ due to age related macular degeneration, cataract, diabetic retinopathy, glaucoma, stroke or other neurological condition, or another cause. A positive response to any of these questions led to the participant being coded as having an eye disease (binary coding variable: presence/absence), except where the only disease reported was cataract, they reported having had surgery and they were not registered as sight impaired.

### Mental ill-health

Mental ill-health was defined when the code ‘mental illness/depression/anxiety/nerves’ was used at least once by the interviewer in response to the question about long-term health conditions described above (binary coding variable: presence/absence).

### Mental wellbeing

At the second stage of data collection, participants over 16 years of age were asked to self-complete the Warwick-Edinburgh Mental Well-Being Scale (WEMWS) [23], a 14-item questionnaire to assess positive affect, satisfaction with interpersonal relationships and positive functioning. This instrument has been shown to be unidimensional and to have strong internal consistency, construct validity and reliability [24].

### Statistical analyses

To test (H1) and (H2) a series of linear regression analyses were undertaken. To test (H1) wellbeing was regressed on self-reported vision, with wellbeing as a continuous variable (total WEMWBS score) and self-reported vision as an ordinal variable at five levels (excellent, very good, good, fair and poor). To test (H2), the model was re-run with mental ill-health included as a covariate (categorical variable at two levels), to determine whether the association between vision and wellbeing was caused by depression (multivariate model 1). Next, the model was re-run with eye disease included (categorical variable at two levels) to determine whether any association was only caused by having an eye disease (multivariate model 2). Finally, the model was re-run with mental ill-health, eye disease, age, sex, socioeconomic status and ethnic group (White British or other) added simultaneously (multivariate model 3). Post-hoc analyses using Student’s t-test pairwise comparisons explored differences in wellbeing associated with each level of self-reported vision. All analyses were performed in JMP (Pro v18.0.2, JMP Statistical Discovery LLC, Cary, NC, USA).

### Patient and public involvement

This analysis was motivated by author MDC’s clinical observations of reduced wellbeing in people with mild vision impairment. The importance of the research was determined by discussion with representatives from patient groups (primarily Stargardt’s Connected and the Macular Society). Dissemination to patient communities has been performed at the Moorfields Biomedical Research Centre ‘Coffee Hour’ by author MDC and is planned for further patient groups.

### Ethical approval

The Health Survey for England data are anonymised and publicly available; as such, no ethical approval or consent was required for these secondary analyses.

## Findings

Data were available for 10,980 participants, of whom 8,795 were over 16 years of age. Mental wellbeing data were not available for 1,069 adult participants: 266 participants chose not to complete this questionnaire, five answered ‘don’t know,’ and for 798 data were scored as ‘not applicable,’ largely due to the nurse visit not being completed. Self-reported vision status was not available for a further 21 people. Data from the remaining 7,705 participants were used for subsequent complete case analyses. This final sample included 3,391 men and 4,313 women, with mean age 49.5 years (sd: 18.3; range 16-104).

Self-reported vision was associated with poorer mental wellbeing (*F* _(4,7700)_ = 94.7, p < 0.0001, Figure 1) and explained 5% of the variance in the outcome variable (*R*^2^ = 0.047). Relative to reporting ‘poor’ vision, each subsequent level of vision predicted better wellbeing, with the exception of ‘fair’ vision, which was not significantly different (Table 1).

**Figure 1.**
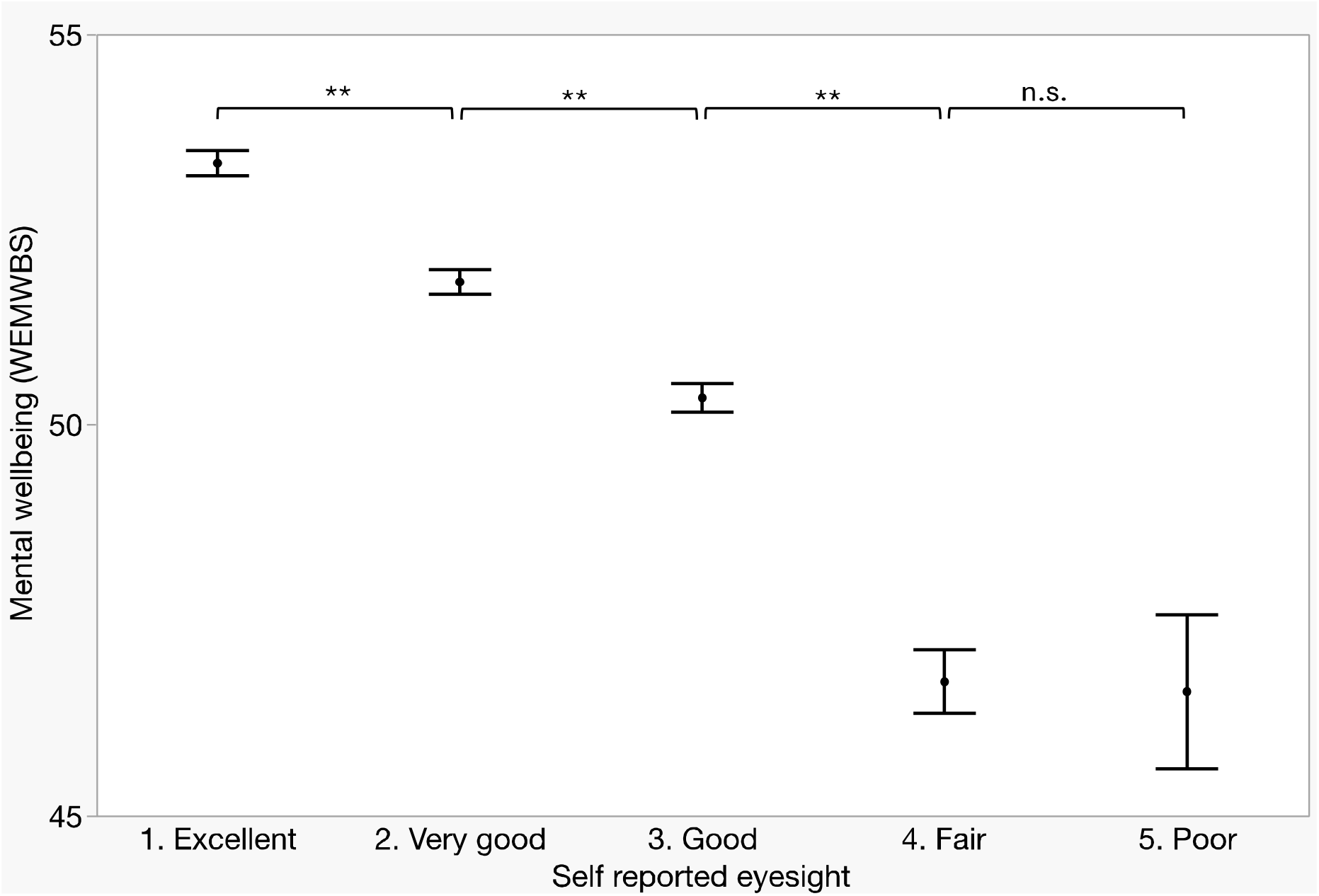
The relationship between self-reported vision and mental wellbeing. Points represent mean values. Error bars show standard error of the mean. Dashed line separates those defined as having vision impairment from those without. ** = significant at *p* < 0.0001 (Student’s t-test). n.s. = not significant, *p* > 0.05.

**Table 1.** Regression analyses showing the regression of wellbeing on self-reported vision, mental ill-health, eye disease, demographic and socioeconomic variables. Values in bold indicate significant predictors. Reference levels for categorical predictors were (in brackets) as follows: self-reported vision (poor), mental ill-health (yes), eye disease (yes), gender (male), ethnicity (white British), socioeconomic status (routine or manual).

|  |  | Univariate models |  | Multivariate model 1 |  | Multivariate model 2 |  | Multivariate model 3 |  |
| --- | --- | --- | --- | --- | --- | --- | --- | --- | --- |
| Predictor | Level | Coefficient<br>(95% CI) | p value | Coefficient<br>(95% CI) | p value | Coefficient<br>(95% CI) | p value | Coefficient<br>(95% CI) | p value |
| Self-reported vision | Excellent | **6.05**<br>**(4.57, 7.53)** | **<0.0001** | **6.05**<br>**(4.57, 7.53)** | **<0.0001** | **6.81**<br>**(5.25, 8.38)** | **<0.0001** | **5.66**<br>**(4.17, 7.16)** | **<0.0001** |
|  | Very good | **4.49**<br>**(3.01, 5.98)** | **<0.0001** | **4.49**<br>**(3.01, 5.98)** | **<0.0001** | **5.29**<br>**(3.73, 6.85)** | **<0.0001** | **4.04**<br>**(2.55, 5.53)** | **<0.0001** |
|  | Good | **3.12**<br>**(1.64, 4.60)** | **<0.0001** | **3.12**<br>**(1.64, 4.60)** | **<0.0001** | **3.80**<br>**(2.24, 5.35)** | **<0.0001** | **2.71**<br>**(1.23, 4.19)** | **0.0003** |
|  | Fair | 0.01<br>(-1.59, 1.61) | 0.99 | 0.01<br>(-1.59, 1.61) | 0.99 | 0.15<br>(-1.52, 1.83) | 0.86 | -0.24<br>(-1.83, 1.35) | 0.77 |
| Mental ill-health | No | **10.6**<br>**(9.82, 11.4)** | **<0.0001** | **10.2**<br>**(9.40, 10.9)** | **<0.0001** | - | - | **9.73**<br>**(8.97, 10.5)** | **<0.0001** |
| Eye disease | No | **1.11**<br>**(0.59, 1.63)** | **<0.0001** | - | - | 0.10<br>(-0.42, 0.62) | 0.70 | **0.61**<br>**(0.07, 1.15)** | **0.028** |
| Sex | Female | -0.30<br>(-0.68, 0.08) | 0.13 | - | - | - | - | 0.08<br>(-0.43, 0.28) | 0.67 |
| Age |  | 0.00003<br>(-0.01, 0.01) | 0.95 | - | - | - | - | **0.03**<br>**(0.02, 0.04)** | **<0.0001** |
| Ethnicity | Other | **1.28**<br>**(0.76, 1.80)** | **<0.0001** | - | - | - | - | **1.14**<br>**(0.64, 1.64)** | **<0.0001** |
| Socioeconomic status | Higher managerial or professional | **2.84**<br>**(2.41, 3.27)** | **<0.0001** | - | - | - | - | **2.13**<br>**(1.72, 2.54)** | **<0.0001** |
|  | Intermediate | **1.07**<br>**(0.56, 1.58)** | **<0.0001** | - | - | - | - | **0.67**<br>**(0.20, 1.15)** | **0.0058** |
|  | Other | 0.097<br>(-1.08, 1.28) | 0.87 | - | - | - | - | 0.07<br>(-1.05, 1.19) | 0.90 |

This association remained significant when mental ill-health (*F* _(4,7699)_ = 87.3, p < 0.0001, multivariate model 1) and eye disease (*F* _(4,7699)_ = 90.2, p < 0.0001, multivariate model 2) were added (separately) as covariates. Further, the relationship remained significant after sex, age, ethnic group and socioeconomic group were added (simultaneously) as further covariates alongside mental ill-health and eye disease (*F* _(4,7681)_ = 76.6, p < 0.0001, *R*^2^ = 0.14, multivariate model 3).

## Discussion

In support of hypotheses H1, we have shown a robust association between self-reported vision and mental wellbeing. Further, consistent with H2, this relationship remained significant after controlling for mental ill-health and the presence of eye disease, as well as a number of socioeconomic and demographic covariates (age, sex, ethnic group and socioeconomic status). Our analyses did not show a difference in wellbeing between those with ‘fair’ vision and those reporting ‘poor’ vision. Self-reported ‘fair’ or ‘poor’ vision has been shown to be sensitive and specific for defining vision impairment in analyses of data from older adults in three studies (the Irish Longitudinal Study on Ageing, the MEC Assessment and Management of Older People in the Community trial and the Health and Retirement Survey) [25,26]. This suggests a plateau effect in our data: progressively poorer vision correlates with poorer wellbeing until the level of ‘vision impairment’ is reached, after which changes in vision are not associated with further lowering of wellbeing scores.

Strengths of this study include its large, representative and wide-ranging population of participants, its use of a reliable and well-validated instrument to assess wellbeing, and our inclusion of multiple potential confounders in our statistical modelling. There are three key weaknesses. First, we used self-reported measures of vision and mental health status. Although there is a precedent for using self-reported vision status in population studies [3,5,27] this may lead to a small bias towards overreporting [25,26] or underreporting vision impairment [28]. We also relied on self-report of mental ill-health and eye disease, the results of which may have been limited by recall bias, by not reporting undiagnosed mental health conditions, or by self-diagnosis of mental health conditions without diagnostic criteria being met. Second, the cross-sectional nature of the study precludes our ability to determine causality or the direction of causality. For example, it is possible that people with reduced wellbeing tend to underestimate how good their vision is, although depression itself is not generally associated with inaccurate visual acuity testing.[29] A previous longitudinal study has shown a bidirectional relationship between anxiety and vision impairment, where people with vision impairment experienced greater anxiety, but people with anxiety were also more likely to become vision impaired, perhaps due to late presentation to services, poor lifestyle factors (such as diet), or their anxiety being caused by having a vision-threatening condition.[27,28] Regardless of the direction of association in our study (poor vision leading to poor wellbeing, or poor wellbeing leading to poor reported vision), interventions to improve vision are likely to improve wellbeing. A final possible limitation relates to data collection: the WEMWS was self-completed by participants as part of a booklet, which is likely to have made it less accessible to people with vision impairment. This is supported by the fact that wellbeing data were available for 81.3% of those with ‘fair’ or ‘poor’ vision, compared to 88.6% of those reporting better vision, a statistically significant difference (Pearson χ2 = 35.7, p < 0.0001).

Our findings build on existing research into wellbeing in people with vision impairment [3,4], and show that even among those *without* vision impairment, better self-reported vision is associated with better mental wellbeing. This finding agrees with Mojon-Azzi and colleagues, who also found an association between self-reported vision and certain aspects of wellbeing in an older adult sample. Our study supports and extends these findings to a much wider age range and uses a validated and more rigorous instrument to assess wellbeing (the WEMWS). Our results also reflect the findings of Cumberland et al, who used UK Biobank data to show that mildly reduced vision was associated with increased mental ill-health and poorer self-rated general health.[6] This work further extends previous research by showing that the effects persist after controlling for a number of potential key confounders. For example, the association between self-reported vision and wellbeing applied to those with and *without* a diagnosis of mental ill-health. This suggests that, consistent with stepped care and early intervention models of intervention, people with vision impairment or who report reduced vision may benefit from supports to improve their wellbeing, even if they do not have a diagnosis of depression or any other mental health condition. Our findings are consistent with calls for the integration of mental and physical health care, and the provision of mental health support in physical health services.[30]

The reduced wellbeing we have identified in people with poorer vision may represent subclinical depression [31] or may reflect people who have depression but have not been formally diagnosed. Nollett and colleagues have shown that many people with vision impairment display signs of depression but are not receiving treatment.[32] Early mental health intervention has been suggested as an important component of vision rehabilitation,[33] although the evidence base for psychosocial interventions improving the mental health of people with vision impairment remains limited.[34] Incorporating mental health support within eye clinics may support positive mental health outcomes.[35,36] The fact that wellbeing is reduced even in people who rate their vision as ‘very good’ or ‘good’ relative to ‘excellent’ suggests that wellbeing may be increased by early treatment of eye disease, for example surgery for mild cataract, which might improve vision from ‘good’ to ‘very good’. People without eye disease also showed an association between self-reported vision and mental wellbeing. This may reflect undiagnosed eye disease, or the need for an updated refractive correction in people who need spectacles but do not have an eye disease.

Future research should adopt a longitudinal design to determine whether ophthalmological intervention in early eye disease does indeed improve wellbeing and whether psychological treatments such as stepped care,[37] intended to improve wellbeing, can prevent the subsequent development of depression in people with vision impairment. This would support calls for integrating mental and physical health services.[30,36] The wellbeing of people from lower socioeconomic groups, who are known to have poorer access to eyecare services,[38] also merits further research.

## Data Availability

Data is publicly available

https://doi.org/10.5255/UKDA-SN-7649-2

## Notes

### Competing Interest Statement

The authors have declared no competing interest.

